# Scoping review of Structured Diagnostic Interviews in the Forensic Psychiatric Setting

**DOI:** 10.1101/2024.06.18.24308925

**Authors:** Jade C Bouwer, Taryn Williams, Karen Marè, Nyameka Dyakalashe, Dan J Stein

**Author notes:** **Correspondence:** Jade Bouwer, Department of Psychiatry and Mental Health, University of Cape Town, Valkenberg Hospital, Observatory Road, Observatory, 7395, Cape Town, South Africa. **Funding:** This research did not receive any specific grant from funding agencies in the public, commercial, or not-for-profit sectors.

## Abstract

**Background:** Structured clinical diagnostic interviews are widely used in clinical practice and psychiatry research. Nevertheless, the extent to which such interviews have been used in forensic psychiatry is unclear, perhaps because of concerns about feasibility and utility.

**Aim:** We undertook a scoping review to investigate publications on structured clinical interviews in the forensic psychiatry context, paying particular attention to issues of feasibility and utility.

**Methods:** A PubMed and PsychInfo database search was undertaken using the terms “structured diagnostic interviews” AND “forensic psychiatry” AND “clinical attitudes” OR “utility” OR “feasibility” OR “acceptability”. PRISMA extension for Scoping Reviews (PRISMA-ScR) was used as a guideline in reviewing and including studies.

**Results:** We found three articles on the use of structured diagnostic interviews in the forensic psychiatry context. In most publications, these interviews were used to assess the accuracy of symptom measures using existing validation tools. There were no publications that reported on issues of feasibility and utility.

**Conclusions:** Literature on the use of structured diagnostic interviews in forensic psychiatry is sparse. While this may reflect concerns about feasibility and utility, no publications provide data on the feasibility and utility of such interviews in the forensic setting. This highlights an important area of research to explore.

## 1. Introduction

Modern psychiatric nosology is governed by two major classification systems (Clark et al., 2017). Currently, mental disorders are diagnosed using diagnostic criteria outlined in the American Psychiatric Association (APA)’s Diagnostic and Statistical Manual (DSM) or diagnostic guidelines outlined by the World Health Organisation’s (WHO) International Classification of Diseases (ICD) 11^th^ edition (ICD-11) (American Psychiatric Association, 2022; First et al., 2015; World Health Organisation (WHO), 2019). Recent revisions of these systems were advised by the nosological evidence-base and aim to inform psychiatric practice and research.

Structured diagnostic interviews (SDIs) for mental disorders are widely used in clinical practice and in psychiatric research. These interviews may improve diagnostic reliability (Basco et al., 2000; Endicott, 2001; Miller et al., 2001; Rocha Neto et al., 2023; Shear et al., 2000). Additionally, their use may promote detection of under-recognised comorbidity (Basco et al., 2000; Pinninti et al., 2003; Zimmerman & Mattia, 1999).

Forensic psychiatric interviews pose unique challenges to the clinician (Logan, 2018). Information obtained in such contexts is not solely for the purpose of diagnosis and treatment implementation but may very well influence court recommendations. These interviews are no longer for the benefit of the patient, but instead serve a third-party client (the court or lawyer) thus opening itself for scrutiny and query. The findings of these interviews thus bear legal weight, and clinicians may be called to account for their assessment procedure and veracity of information obtained. The availability of biological testing would certainly appease the courts in the reliability and sensitivity of information collated (Endicott, 2001), eliminating errors not only on the part of the clinician but also the interviewee. Such a luxury is not afforded to the field of psychiatry. By systematically probing symptoms and behaviours, variability between interviewers may be reduced (Basco et al., 2000) and dynamic factors such as rapport and clinician expertise accounted for.

Nevertheless, the extent to which such interviews have been used in forensic psychiatry is unclear. Our anecdotal impression is that they are not widely employed. Barriers to their use might include concerns about feasibility and utility. For example, interviews are often lengthy and require training (Basco et al., 2000), and existing interviews may not always address diagnoses that are key in the forensic setting (e.g., antisocial personality disorder, intermittent explosive disorder).

We undertook a scoping review to investigate publications on structured clinical interviews in the forensic psychiatry context, paying particular attention to issues of feasibility and utility.

We aimed to (1) search for all published English-language studies on structured diagnostic interviews undertaken in the forensic psychiatric setting, (2) determine which SDIs have been used in these studies, and (3) collate any data on clinician attitudes, utility, feasibility, and acceptability of SDIs in the forensic setting.

## 2. Methods

The undertaking of this review was guided by The PRISMA extension for Scoping Reviews (PRISMA-ScR) (Tricco et al., 2018). A proposal of this scoping review was submitted to a panel of experts in the field and approved.

### 2.1 Search methods

PubMed and PsycINFO were searched for studies on SDIs in the forensic psychiatric setting. Only studies published in English and dated post 1980-Feb 2024 were included. Broad terms such as “structured diagnostic interviews” AND “forensic psychiatry” AND “clinical attitudes” OR “utility” OR “feasibility” OR “acceptability” were used when searching for included studies. Search terms were limited to title and abstract to yield the most relevant results.

### 2.2 Criteria for considering and including studies

Only studies utilising structured diagnostic interview tools on adult (older than 18) psychiatric patients admitted to forensic psychiatric units were included. These patients could be undergoing court-appointed evaluation or have already been declared forensic psychiatric patients by the courts post-assessment. The included studies could be observational or comparative studies, but had to comment on the clinically utility, feasibility, or acceptability of the use of these tools in this population.

### 2.3 Data collection, extraction, and management

All papers sourced were exported to Endnote™ (The EndNote Team, 2013), an online referencing manager software package. Once duplicates were removed the studies were further exported to Rayyan (Ouzzani, Hammady & Fedorowicz, 2016) and screened by two independent reviewers. First, the two reviewers reviewed the abstracts and titles of the studies for inclusion. Second, reviewing the selected studies full texts for inclusion. Any disagreements in selecting and making final decisions for including studies in the review were resolved by means of reviewer discussion (Figure A.1). Since this was a scoping review, articles were not excluded based on quality.

Once the final selection of included studies was made, select data was extracted: 1) authorship, year, 2) the name of tool, 3) setting, 4) sample studied and size, 5) rationale for use of the tool, and 6) any information gathered about the tool (e.g., feasibility and utility). These data were then synthesised into a table (Table A.1).

### 2.4 Data analysis

A narrative analysis was done which included the description of the studies and findings found specific to the objectives of the review. Since this was a scoping review, a quality analysis was not conducted on these studies.

## 3. Results

Our primary search yielded 121 unique articles. 99 articles were excluded based on title and abstract review. 22 full-text articles were reviewed. Out of the 22 full-text articles only three studies were found that met the inclusion criteria of the review and were analysed (Figure 1). Findings of these articles are represented in Table 1 and a narrative study provided below. Studies that were excluded were mostly because of wrong outcomes (82), wrong population (67), wrong study design or duration (47 + 1) or for being in a foreign language and therefore not analysed (3).

Two of these studies were published in the last decade (Green et al., 2013; Tylicki et al., 2018), and one in 2004 (Kristiansson et al., 2004). All studies were conducted at forensic psychiatric sites in high-income countries (HICs) (America and Sweden) although one included a migrant population living in an HIC (Kristiansson et al., 2004). All three studies were conducted on defendants undergoing a forensic psychiatric evaluation (FPE). Additionally, one study included a simulation group (Green et al., 2013) and one included civil litigation referees and post-trial participants (Tylicki et al., 2018). Most participants in all three studies were male between the ages of 30 – 40 years. Psychiatrists and psychologists conducted the studies in all three articles; however, two studies (Kristiansson et al., 2004; Tylicki et al., 2018) also included a self-report component. Studies ranged from 50 to 478 participants. Psychotic disorders tended to be the most common diagnosis among participants (Green et al., 2013; Kristiansson et al., 2004) and the most common offences triggering the FPE involved violent offences such as sexual or physical assault (Kristiansson et al., 2004; Tylicki et al., 2018).

Two studies evaluated the accuracy of the Structured Interview of Reported Symptoms, Second Edition (SIRS-2) in comparison to the original SIRS (Green et al., 2013; Tylicki et al., 2018). The SIRS is a structured diagnostic interview tool which is considered the gold-standard for the assessment of feigning of symptoms of mental illness (Green et al., 2013). The first of these, a study by Green, Rosenfield and Belfi (2003) (Green et al., 2013) aimed to determine the sensitivity and specificity rates for both, and then determine the additive value of the RS-Total scale and the MT index of the SIRS-2. To do this, the sample studied included defendants undergoing criminal forensic evaluation and a community simulation sample. The second study to look at utility of the SIRS was undertaken by Tylicki et al. (2018) (Tylicki et al., 2018). It sought to examine the associations of the SIRS and SIRS-2 by comparing the classification scales to those of the Minnesota Multiphasic Personality Inventory-2-Restructured Form (MMPI-2-RF) validity scales.

Both these studies reached a similar conclusion regarding the clinical utility of the updated SIRS-2. Green, Rosenfield and Belfi (2003) (Green et al., 2013) found the specificity of both in the forensic sample to be slightly improved (94.3% vs 92%) but sensitivity reduced (36.8% vs 47.4%) (Table 1). Similarly, Tylicki et al. (2018) (Tylicki et al., 2018) found the specificity of the SIRS-2 to be 100% in both the civil and forensic samples (in comparison to the SIRS which had a specificity of 100% and respectively), but a reduced sensitivity (9.3% in the civil litigants and 41.2% in the forensic sample). Therefore, the ability of the SIRS-2 in identifying false positives may be increased, the sensitivity was greatly reduced.

Furthermore, the addition of the RS-Total scale did not have any beneficial outcome on improving sensitivity (Green et al., 2013). Both studies also then explored the utility of the classification measures of the SIRS-2 and established that it held limited value in correctly identifying feigners from indeterminate groups as assigned by the original SIRS (Green et al., 2013; Tylicki et al., 2018) with a recommendation to review cutoff limits.

Further, Kristianson, Sumelius and Sondergaard (2004) (Kristiansson et al., 2004) looked at the feasibility of using the Clinician-Administered PTSD Scale (CAPS) versus the Structured Clinical Interview for DSM-IV (SCID) – PTSD, both structured questionnaires, to identify post-traumatic stress disorder (PTSD) in FPEs. A cohort of Swedes were compared to an immigrant group with similar criminal profiles. The SCID-PTSD was found to be less sensitive in immigrants, raising a question regarding its utility in interpreting symptoms in non-Swede speakers. The CAPS was therefore found to be more clinically adept at diagnosing PTSD across heterogenous groups, and clinicians found it to be more accurate and structured.

## 4. Discussion

We found 3 articles on the use of SDIs in the forensic psychiatry context. In most of the publications, these interviews were used to assess the accuracy of symptom measures themselves rather than the use/purpose of these tools in this population. While this is beneficial in aiding a clinicians’ decision in which tool to consider and how reliable these may be, they still do not answer the question of attitudes or acceptability. The studies also did not comment on the time taken to conduct each interview, leaving open-ended questions regarding their feasibility in the context of human resources and time.

These findings conflict with the use of SDIs in other settings. Randomized controlled trials frequently use these instruments to ensure inclusion/exclusion criteria are met. Studies of the neurobiology of mental disorders similarly often use these instruments to characterize their samples, although there has been a move to use other assessment approaches e.g., Research Domain Criteria (RDoC) (Morris et al., 2022). The infrequent use of these tools in a forensic setting not only lends itself to the fallibility of the clinical interview (subject to a range of variables such as clinician experience and opinion, therapeutic rapport, and patient recall) and diagnostic inaccuracy which may bear weight on treatment outcomes but allows for legal scrutiny of the clinician compiling such reports as to their scientific, evidence-based justification for clinical opinions expressed (Logan, 2018).

Several limitations of this scoping review deserve emphasis. First, we did not assess papers published in languages other than English. However, during our search, we came across three such papers. Second, we did not assess the grey literature. However, given how sparse the literature in formal databases is, we do not expect that there is a large grey literature. Third, only two databases were searched, and the search terms used highly specific to enhance accuracy of the database search; articles not listing any of the keywords used but of relevance may have been missed. Fourth, the studies were not assessed for quality. Lastly, studies evaluated were all conducted in HICs and may not be generalisable to settings with high patient to clinician ratios, overburdened forensic services with long waiting times for assessment, and time constraints.

## 5. Conclusion

In summary, the literature on the use of SDIs in forensic psychiatry is sparse. This seems an important gap to address. The legal weight of such assessments demands careful assessment of psychiatric symptomatology. The absence of evidenced-based assessment measures in this setting makes assessing the quality of its findings difficult to assess or defend. Future work in the formulation, implementation and appropriateness of such tools may not only enhance diagnostic accuracy, but also promote uniformity of assessments across sites in this complex population. Not only could this improve clinician satisfaction in their utility and encourage their use, but also build a clinical argument that can stand legal contestation.

## Data Availability

All data produced in the present study are available upon request to the authors.

### 6. Appendices

**Figure A.1:**
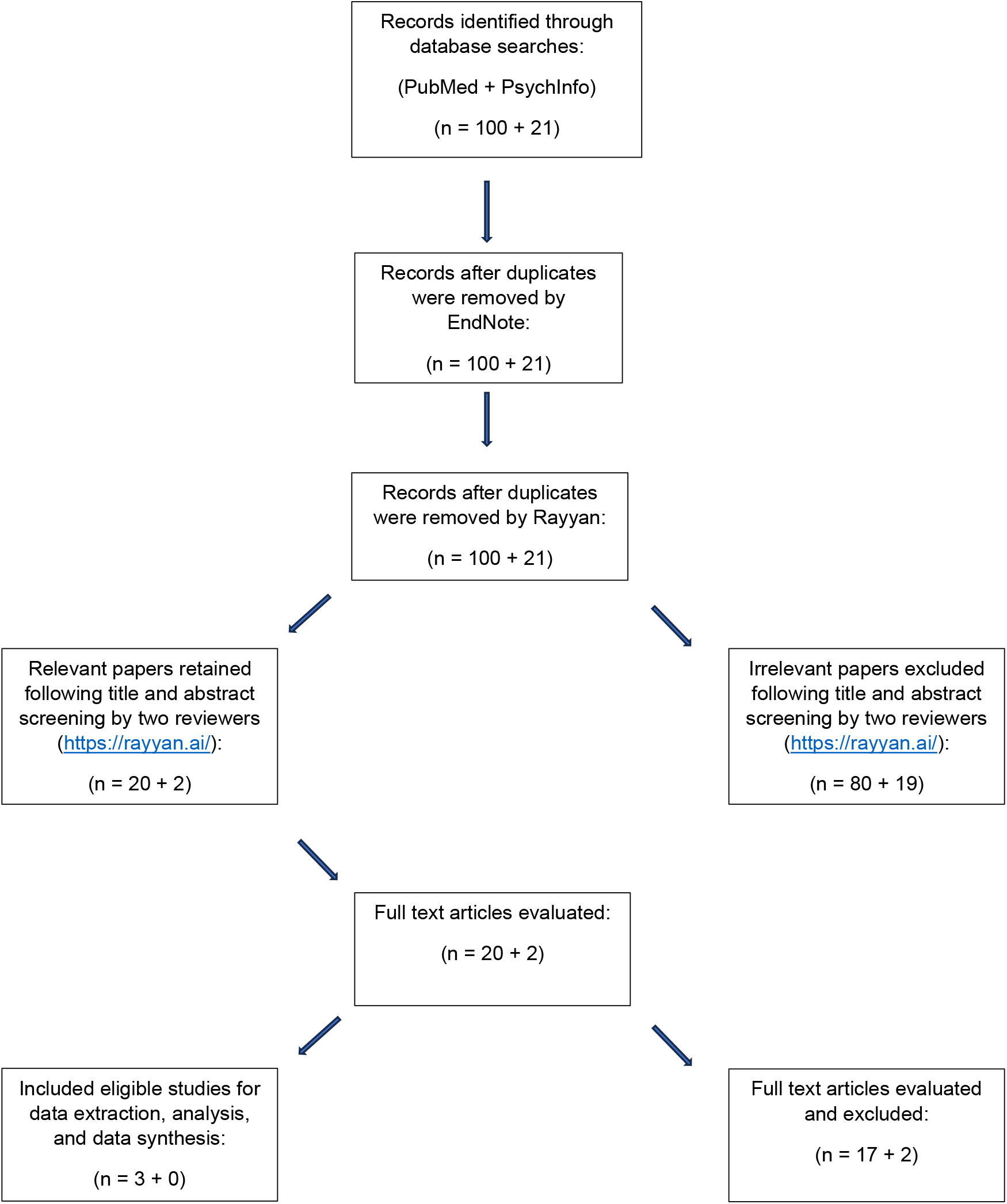
Systematic literature search flow diagram.

**Table A.1:**
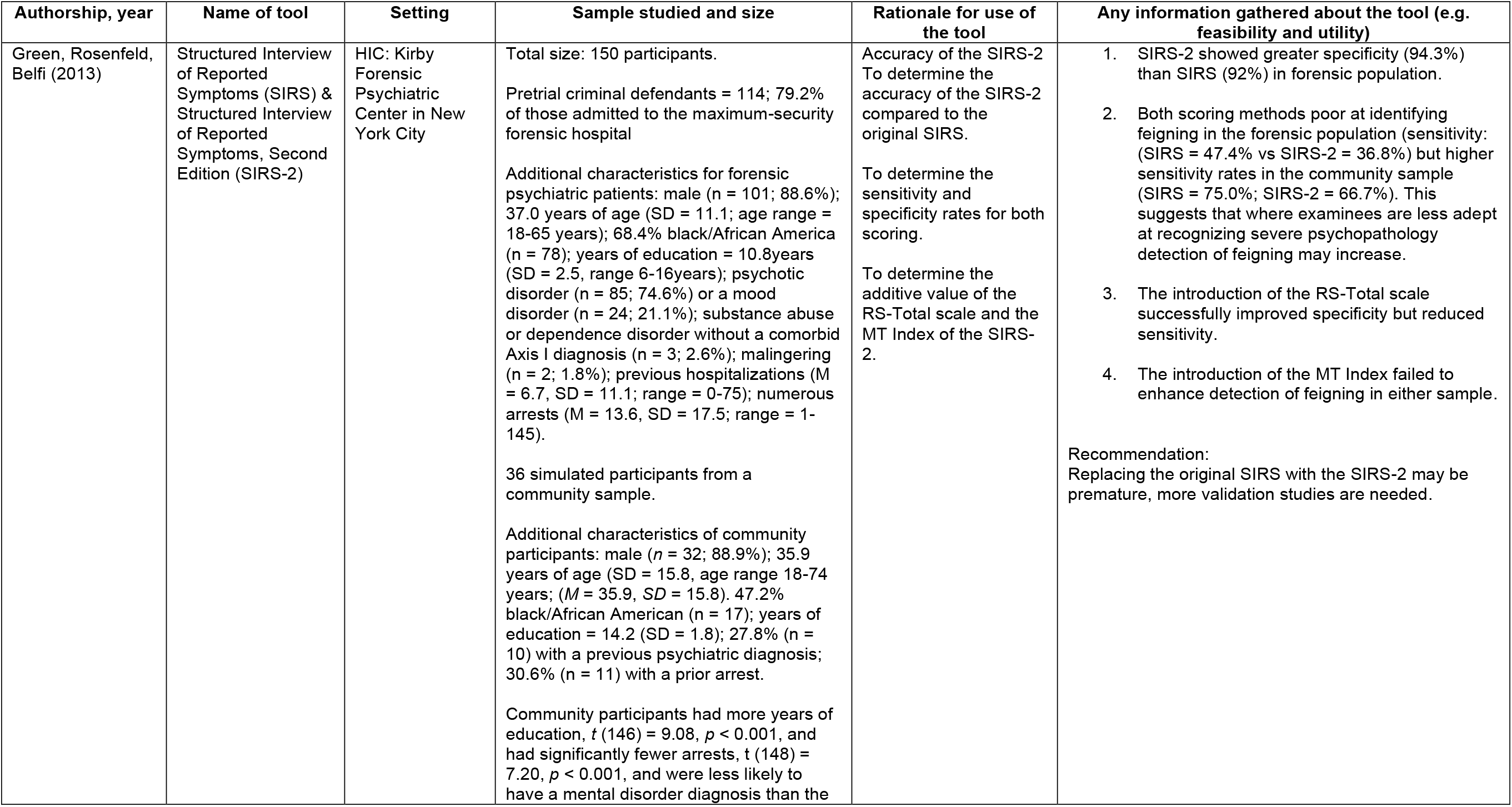
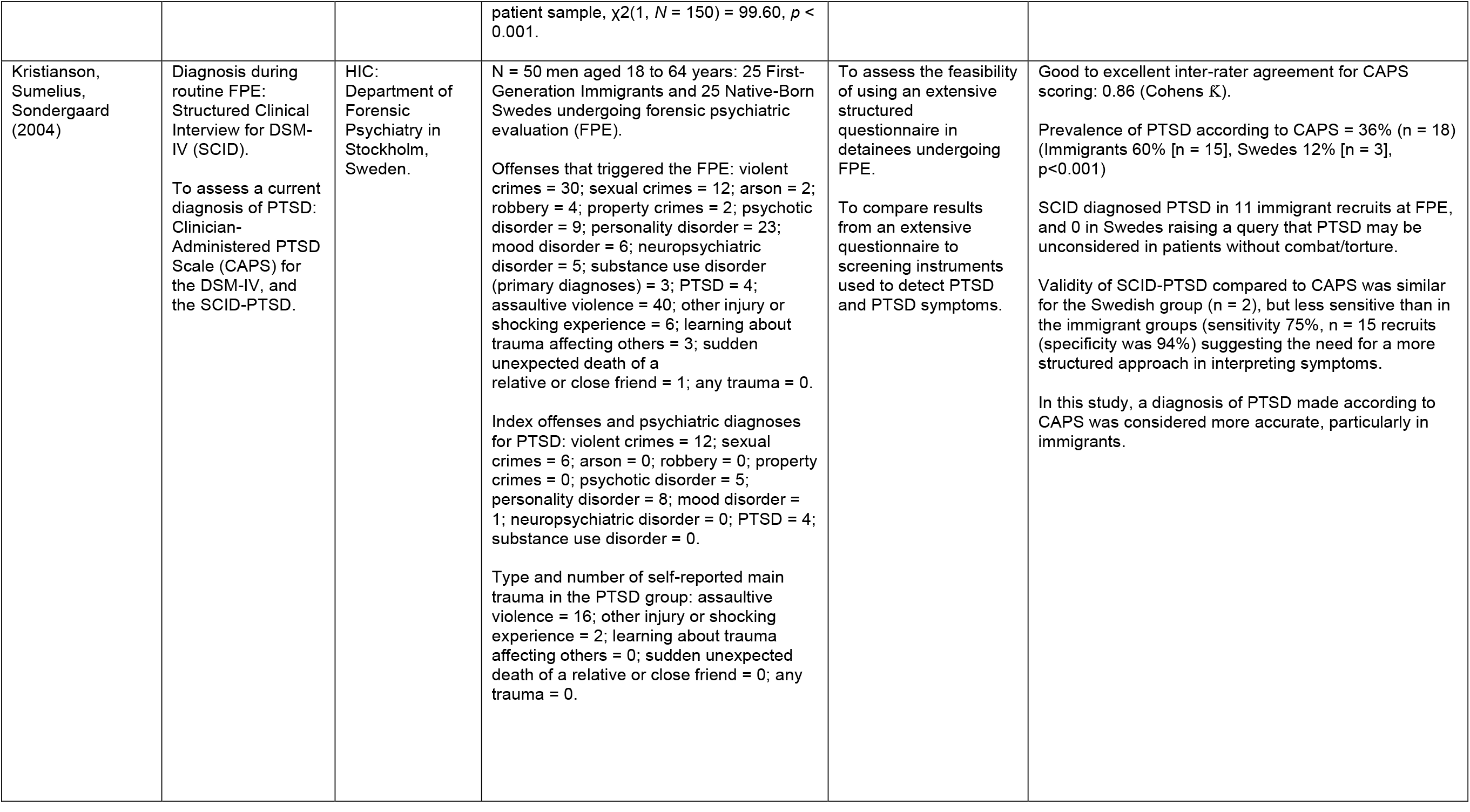
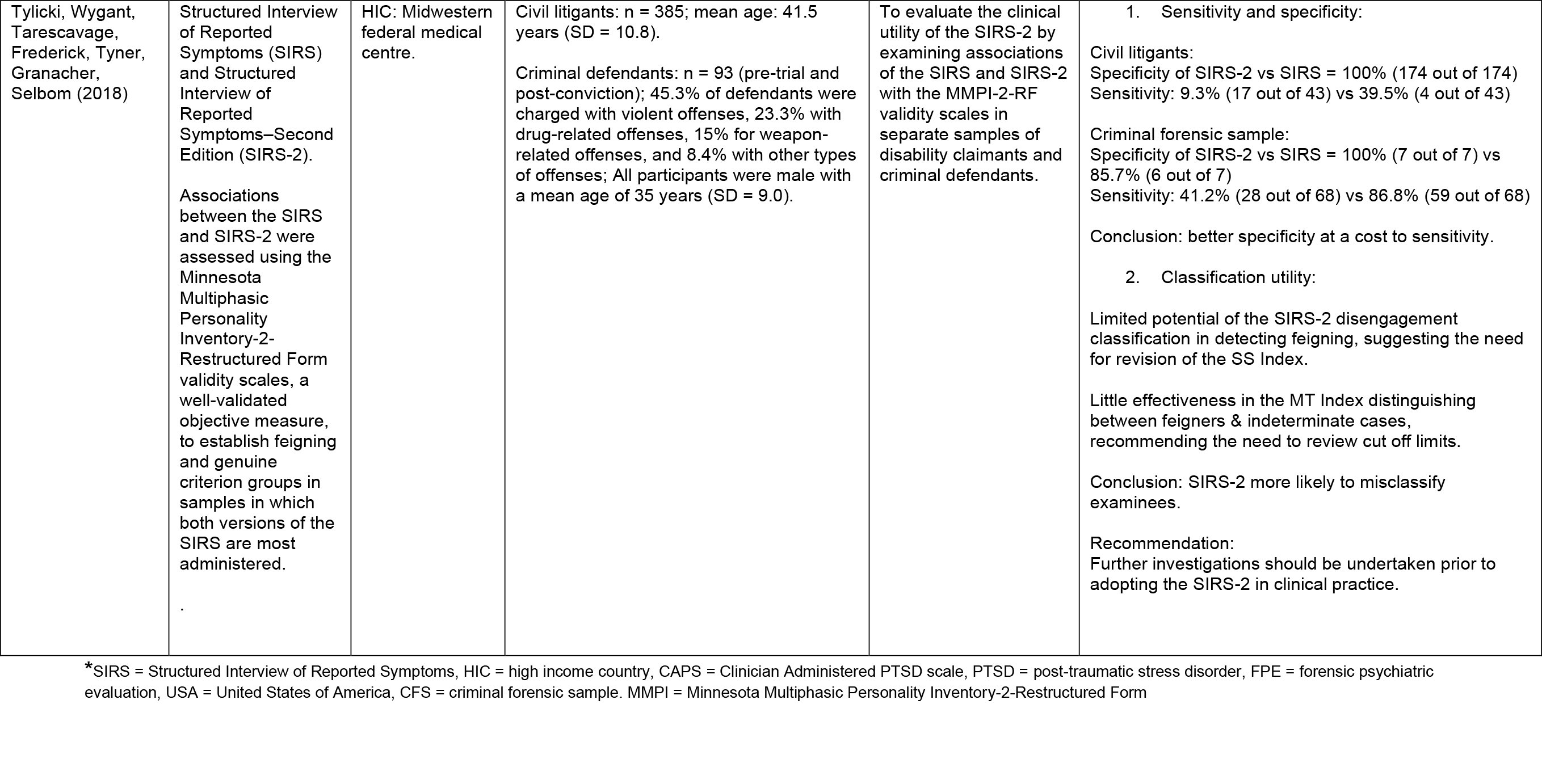
Data Extraction Table of Included Studies.

